# Predicting morbidity by Local Similarities in Multi-Scale Patient Trajectories

**DOI:** 10.1101/2020.09.14.20194464

**Authors:** Lucía A Carrasco-Ribelles, Jose Ramón Pardo-Mas, Salvador Tortajada, Carlos Sáez, Bernardo Valdivieso, Juan M García-Gómez

## Abstract

Healthcare predictive models generally rely on static snapshots of patient information. Patient Trajectories (PTs) model the evolution of patient conditions over time and are a promising source of information for predicting future morbidities. However, PTs are highly heterogeneous among patients in terms of length and content, so only aggregated versions that include the most frequent events have been studied. Further, the use of longitudinal multiscale data such as integrating EHR coded data and laboratory results in PT models is yet to be explored. Our hypothesis is that local similarities on small chunks of PTs can identify similar patients with respect to their future morbidities. The objectives of this work are (1) to develop a methodology to identify local similarities between PTs prior to the occurrence of morbidities to predict these on new query individuals; and (2) to validate this methodology to impute risk of cardiovascular diseases (CVD) in patients with diabetes.

We have proposed a novel formal definition of PTs based on sequences of multi-scale data over time, so each patient has their own PT including every data available in their EHR. Thus, patients do not need to follow partly or completely one pre-defined trajectory built by the most frequent events in a population but having common events with any another patient. A dynamic programming methodology to identify local alignments on PTs for predicting future morbidities is proposed. The proposed methodology for PT definition and the alignment algorithm are generic to be applied on any additional clinical domain. We tested this solution for predicting CVD in patients with diabetes and we achieved a positive predictive value of 0.33, a recall of 0.72 and a specificity of 0.38. Therefore, the proposed solution in the diabetes use case can result of utmost utility to patient screening.

**Highlights:**

- Local similarities between patient trajectories can potentially be used to predict morbid conditions.
- A formal definition of patient trajectories comprising heterogeneous clinical observations, biomedical tests and time gaps is proposed.
- A novel dynamic programming methodology is proposed to find similar patients based on the Smith-Waterman alignment algorithm and a set of customized scoring matrices.

## 1. Introduction

### 1.1. Patient Trajectories

Patient trajectories (PTs) are a proposal for representing the evolution of diseases over time to facilitate their understanding and analysis under a temporal perspective, as well as to discover relationships between patient conditions. The need to use PTs arises due to the complexity of clinical data, which include data from very diverse sources (e.g blood test, images, hospital expenses) and its spread along time. Even though physicians can access this information, usually event by event, on the patients’ Electronic Health Records (EHR), drawing conclusions at a population level under a precision medicine approach becomes a more difficult task. PTs are able to represent the history of a patient as a timeline of every clinical event.

We have found different names for the concept of PT in our research. In [1], 1,171 different *temporal disease trajectories* were defined from the EHR of 6.2 million patients over 15 years using clustering and the Jaccard index as similarity measure. These trajectories compiled the most frequent diagnosis in the development of a disease. Giannoula et al. [2] identified temporal patterns in *patient disease trajectories* using dynamic time warping. They use the concept of distance/dissimilarity between patients to find similar diagnosis codes and build these aggregated trajectories. Both [1] and [2] suggest that the trajectory analysis could be used for the prediction and prevention of disease development, but did not go further on that path. In [3], the frequent process patterns found in *clinical pathways* were used to design time dependency graphs. Given a new patient, they would be assignedto one of those designed pathways. In [4], clustering was used to find 7 frequent *clinical pathways*, according to the encounter types, diagnostics, medications and biochemical measurements of 664 patients. After that, machine learning was used both to assign the patients to one of the 7 created pathways and to predict the next visit of the patient with and without timestamp using only their laboratory results. A very similar strategy was used in [5], were 31 distinct pathways were found from 1,576 patients. In [6] they predict *patient’s trajectory of physiological data* by retrieving patients who display similar trends on their physiological streams, according to the Mahalanobis distance. In this work, they also try to identify which patients will develop Acute Hypotensive Events using these physiological signals.

In this study, we represent patient trajectories as the time-ordered sequences of consultations, laboratory results and diagnosis that each patient has in their EHR. We use PTs to identify partial similarities in patient’s EHR that allow to predict the development of a disease. Patient trajectories are not built according to the most frequent events recorded in EHRs but with all the available information, as aggregating that information could limit the link between patients. Patients do not need to follow partly or completely one pre-defined trajectory, but having common events with another particular patient. In this way, query patients whose EHR includes rare events can also be reflected in the patients in the database, and thus find high similarities during the alignment.

### 1.2. Sequences Alignment

Since a patient trajectory is an ordered sequence of events, the same technology as in biological sequence analysis, such as the alignment of DNA sequences, could be applied to PT analysis. Several well-known bioinformatic algorithms based on dynamic programming allow solving hard alignment problems by splitting the problem into simpler sub-problems. Sequence alignment in bioinformatics aims to identify similar regions in biological sequences under hypotheses of functional, structural or evolutionary relationships.

The alignment can be made i.e. globally, using the Needleman-Wunsch algorithm [7] or locally, using the Smith-Waterman [8]. Both are dynamic programming algorithms, which guarantees finding the optimal alignment according to the scoring system used. Smith-Waterman algorithm (Algorithm 1) performs local alignments of two sequences of symbols of a common alphabet, identifying, as a result, the most similar regions within them. This alignment is done by calculating the Levenshtein distance (or an opposite score) given by three editing operations to transform each pair of symbols (insertion, deletion, or substitution/match), and the possibility to re-start the alignment score from any alignment point (initialization). In consequence, using the Smith-Waterman algorithm for comparing PTs would result in finding high-similar regions between PTs, possibly related to a common disease appearing in the future. This approach may be more adequate than the Needleman-Wunsch algorithm due to the more than likely high heterogeneity of PTs.

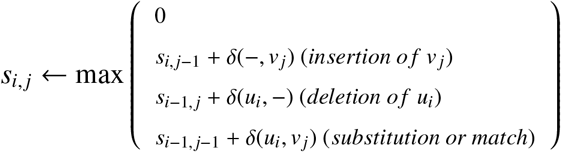

**Algorithm 1** Main instruction of the Smith-Waterman algorithm. The value *δ* of the editing operations consists in a scoring matrix which values change according to the particular use case of the algorithm (e.g homology of proteins, DNA, RNA). In the case of PT comparison, *δ* value is the similarity between EHR events.

Sha et al. work [9] also presented a modified version of the Smith-Waterman algorithm to identify similar patients. They used it to predict mortality in patients with Acute Kidney Injury, based only on their laboratory test data. They did compare the predictive power of their similarity measure against other better known such as the cosine distance and the Jaccard similarity coefficient. They concluded that this Smith-Waterman-based similarity measure achieved better sensitivity and F-measure than the other similarity measures.

### 1.3. Hypothesis

Our hypothesis is that local similarities on small chunks of PTs can identify similar patients with respect to their future morbidities. In other words, we believe that the development of a pathology can be predicted if there is a high local similarity of a PT to a set of PTs of people who developed this pathology. This hypothesis relies on the reasonable assumption that similar patterns in clinical conditions occur in patients during the development of similar disease prognoses. The search and location of these patterns could be used as a screening method in healthy patients.

### 1.4. Use Case: Predict CVD development in Diabetes Mellitus by patient trajectories

In our study, we have tested our hypothesis by assessing the risk of developing cardiovascular diseases (CVDs) in patients with diabetes. Diabetes is a well-known disease with high prevalence worldwide, which is estimated to increase even more by 2045, affecting more than 629 million people in the world [10]. Diabetes causes hyperglycaemia, which results toxic and can cause the development of several health complications, such as ophthalmological, nephrological, neurological and/or cardiovascular diseases. It becomes a priority to diagnose these co-morbidities as soon as possible to improve the patients’ quality of life and reduce economic costs. In this paper, we focus on detecting CVDs as a proof of concept because of the close relationship between cardiopathies and diabetes [11, 12]. This becomes more obvious in the study [12], where they show that while the rate of incidences of myocardial infarction for non-diabetic subjects is 3.5% (18.8% if they have had another infarction previously), in the case of diabetes patients it is 20.2%, (45% if they have had a prior infarction) [13].

## 2. Materials

### 2.1. Dataset

In this study, we used all patients with at least one diagnosis of diabetes mellitus between 2012 and 2015 from Hospital Universitario y Politécnico La Fe, Valencia. Hence, the dataset included 9,670 patients with diabetes mellitus type I or type II, and with or without complications (see Table 1 for details). Each registry consisted of de-identified demographic data (age and gender), timestamped clinical data (diagnostics made in hospitalization or in emergency room), timestamped consultation codes, and timestamped laboratory test results. 425 patients were discarded because they had only one observation on their EHR or they did not have all the necessary identification fields. Hence, from the 9,245 available patients, 3,181 had developed cardiovascular diseases and 6,064 had not. Table 1 also shows the mean and standard deviation of the number of diagnostics, consultations and laboratory test results per patient. It shows how the length of the patient trajectory of people who have developed CVD is larger, due to the development of the disease. It is remarkable that 25% of the patients have less than 10 observations in their trajectory, which means that most of the PTs will contain less information than what it would be expected from a chronic patient (see Figure A.1).

**Table 1:** Exploratory analysis of the dataset. A third of the patients have developed CVD. These patients have more events in their EHR, especially more consultations, therefore longer trajectories.

| | Number of<br>observations | Number of<br>events<br>($\mu \pm \sigma$) | Number of<br>diagnostics<br>($\mu \pm \sigma$) | Number of<br>consultations<br>($\mu \pm \sigma$) | Number of<br>laboratory tests<br>($\mu \pm \sigma$) |
| --- | --- | --- | --- | --- | --- |
| Total | 9670 | 37±38 | 8±7 | 13±21 | 15±17 |
| Used | 9245 | 39±38 | 8±7 | 14±21 | 16±17 |
| With CVD | 3181 | 53±47 | 10±8 | 20±28 | 21±21 |
| Without CVD | 6064 | 31±29 | 6±6 | 10±16 | 13±14 |

### 2.2. Codification

Diagnostics are coded according to ICD-9-CM, which is divided into chapters according to the family of the disease (i.e. diseases related to the circulatory system and CVD belong to chapter 7, diseases related to the genitourinary system makeup chapter 10). A total of 169 consultation and hospital services codes appeared in the dataset, using hospital codes such as CCAR for cardiology and CNEF for nephrology. In addition, some numerical laboratory results have been discretized into ranges such as Low, Normal, and High, according to the thresholds defined by the hospital blood tests.

## 3. Methods

### 3.1. Local Patient Trajectory Alignment (LPTA) algorithm

We have adapted the Smith-Waterman algorithm in order to compare PTs. The computation of PTs comparisons has the following requirements. First, a similarity measure between PTs should be defined. Second, the algorithm should deal with sequences where heterogeneous observations that cannot be compared between them may appear (i.e. laboratory results and diagnosis codes). Finally, the predictive analytics based on PTs should be applied to a massive number of patients. To define a similarity measure between PTs, we establish the next properties:

1. The local similarity measure of one PTs with itself should be maximum.
2. The measure should consider that regions of PTs may contain gaps that do not match. For instance, one patient may have needed more consultations than other between diagnostics during a similar sequence of episodes, and the similarity measure should be able to keep the track of the common events despite of the noise that the extra consultations could add.
3. The similarity measure should penalize differences in time between two consecutive observations.
4. The calculated similarity score will then be used to rank patients of the reference dataset according to their local similarity to any query patient.

The main difference between the classical edit distance of biological sequences, where all the characters represent the same idea (i.e. nucleotides, amino acids), and our PTs similarity measure, is that our sequences may contain observations of different nature. Hence, instead of having a single scoring matrix, as in the original Smith-Waterman problem, we have a set of similarity functions defined between concepts appearing in the PT alphabet (e.g. diagnostics, consultations and laboratory test results):

- The similarity measure between consultations is an indicator function of the consultation services.
- The similarity measure between diagnosis is defined by a combination of indicator functions of categories and subcategories of the ICD-9 codes, weighted by the similarity of locations where the diagnostics were done (emergency room or hospitalization) or by the time relationship with the previous diagnosis.
- For real-valued observations, such as laboratory results, we define similarities of indicator functions after their categorization to have a clear clinical comparison (e.g. both glucose values are in normal or abnormal levels).

These similarity functions will score the similarity amongst the patients not only considering the degree of similarity of the most similar regions between the PTs, but also the similarity of these regions to the typical development of the target disease.

Hence, we define the Local Patient Trajectory Alignment (LPTA) algorithm as a dynamic programming algorithm for finding the most similar regions between PTs (Function 3.1). These regions would be scored according to their direct similarity and their relationship to the development of the disease (e.g. CVD in patients with diabetes mellitus). The Smith-Waterman function of the LPTA procedure works similarly to the original algorithm described in Algorithm 1 but changing how the scoring works: 6 would no longer be a scoring matrix, but a set of scoring functions. A pseudo-code version of the functions involved in the scoring process can be found in the appendix (see Functions Appendix A.1, Appendix A.2), and an explained example of how they work, together with the formal language defined on Section 3.2, can be found in Figure A.2.

**Function 3.1:** LPTA main algorithm. queryPatients is a list of n PTs which condition is wanted to be known, DBPatients is a list of m PTs which condition is already known(LabelDBPatients). queryPatients are aligned to DBPatients using the set of similarity functions DELTA (Appendix A.1) with dMatrices (see Figure3) as parameter. maxScores will store the scores of the alignments between patients.

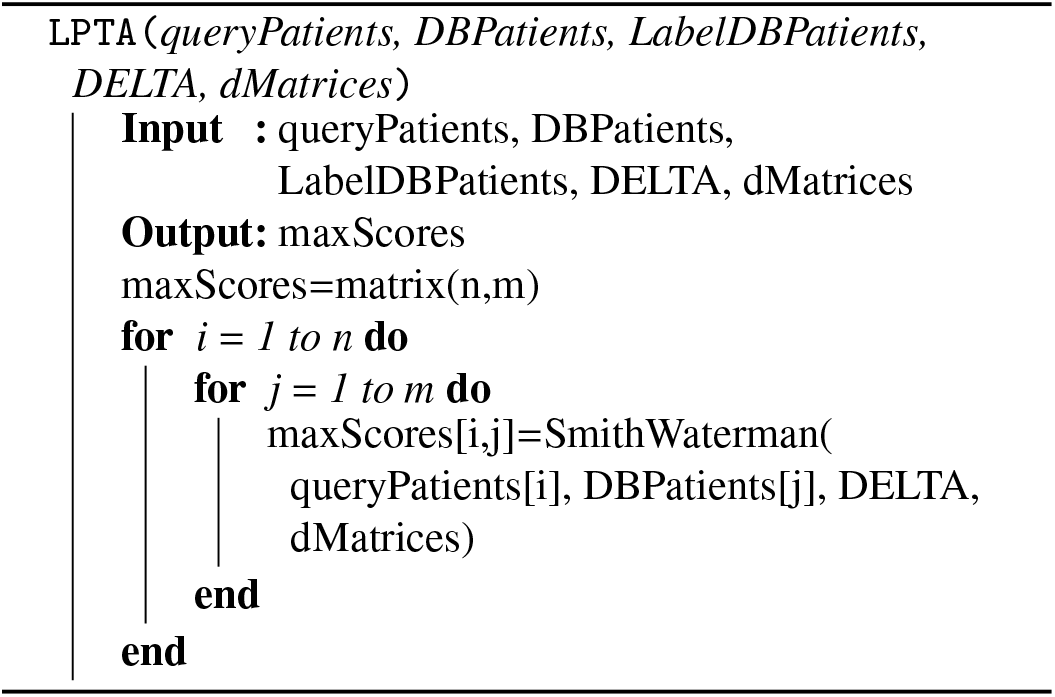

LPTA algorithm returns a vector of scores for each query patient according to its similarity to each PT of the reference database. In order to assign the condition to the query patient based on these scores, a classification method was developed: The query patient would be classified as disease developer if at least one of the N reference patients with a higher similarity score had developed it. N is a parameter to be optimized in the experiments.

It is worth noting that scores are normalized by the length of the reference PT amongst which the query patient is being compared. This way, if the comparisons of a query patient with two reference patients get the same score, it can be assumed that the similarity between the query patient and the patient with fewer observations is higher than similarity to the longer one.

For our experiments, the LPTA algorithm has been implemented using R and the packages [14, 15, 16, 17] for CPU-parallelization, temporal cost calculation and graphical representations. An implementation of the LPTA using Big Data technologies, such as Storm and Redis, is already in development [18]. This will help to decrease the temporal cost of the algorithm, allowing us to analyse massive amounts of PTs for screening parallelly query patients. This is the desired real use of the LPTA.

### 3.2. Patient Trajectory Formal Definition

We propose a formal language for defining patient trajectories from EHR data and computing local similarities using the proposed LPTA algorithm (Function 3.1). In this section, we define the formal language of patient trajectories for our use case, but this grammar could be easily adapted to another problem’s needs. Every event included in the EHR that had every field needed (consultation type, diagnosis code, timestamp, etc.) will be included in the PT. If any of these fields was missing, the event would not been added in the PT.

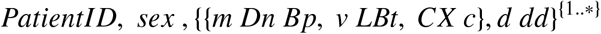

The PT definition can be found in (2), where *PatientID* is the identifier of the patient, *sex* is the sex of the patient (F if female or M if male), *m* is an ICD-9 code, *n* can be either H if the diagnosis was made in hospitalization or E if it was made in emergency room, *p* can be either E if the diagnosis is related to a previous emergency or C if not, *v* is a numerical result of the laboratory test, *t* is the laboratory test type (i.e. T for total cholesterol, H for HDL, C for creatinine and L for glycosylated haemoglobin) and *c* a consultation code. In addition, *d* is the number of days from the previous event, whichever its type is, whereas *dd* is the number of days from the very first event recorded in the EHR. The first temporal parameter reports the relationship between the episodes and the second one the density of observations. The greater the density, the more times the patient would have been to the hospital and the greater the chances that they are developing a pathology. These two parameters avoid having to work with timestamps. Two explained instances of this formal language are shown in Figure 1 and Figure A.2.

**Figure 1:**
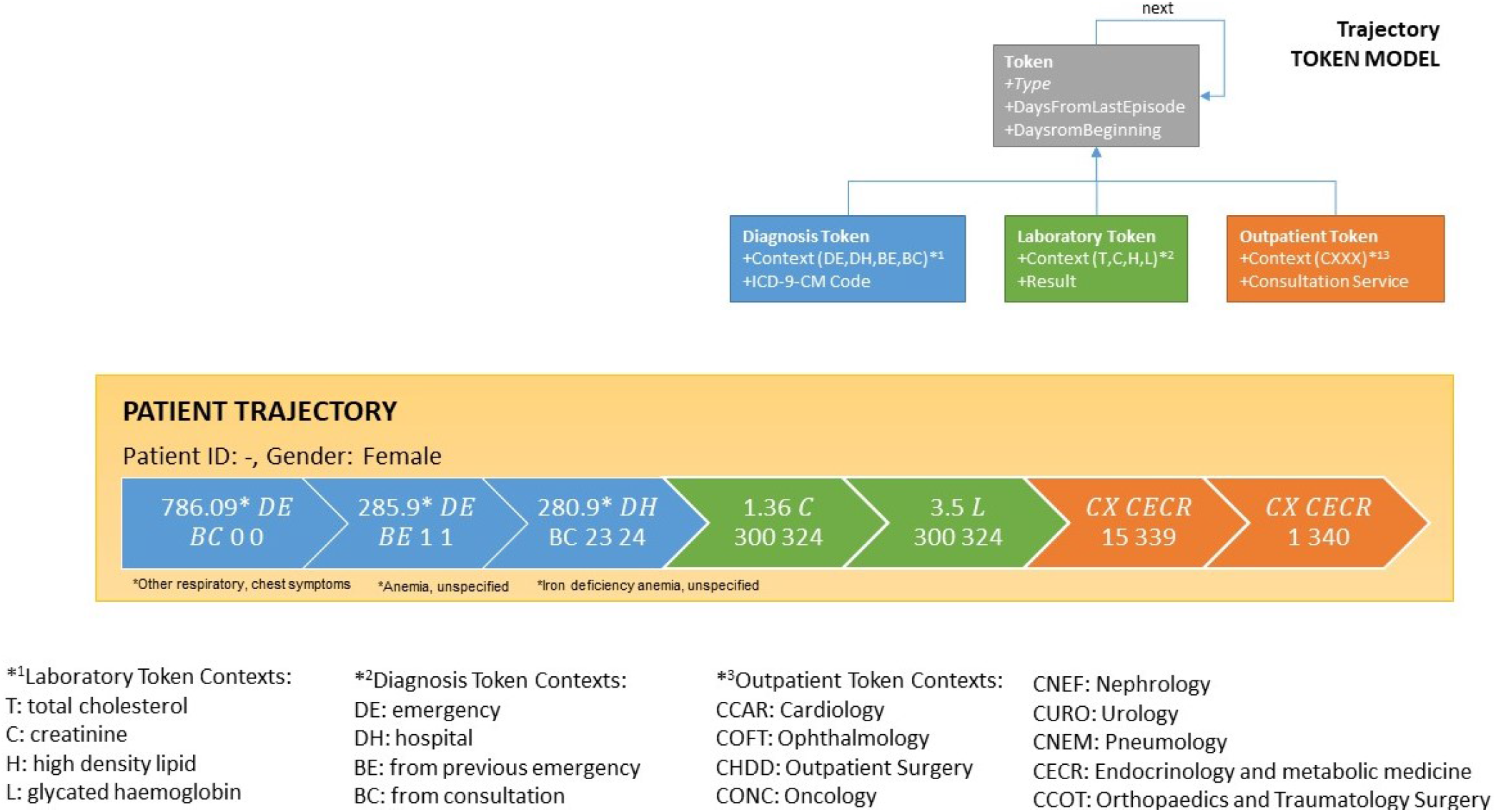
An example instance for a patient trajectory and the trajectory token model. Three diagnostics events can be seen, followed by two laboratory results and two consultations. The PT would be: -, F, 786.09 DE BC 0 0, 285.9 DE BE 1 1, 280.9 DH BC 23 24, 1.36 C 300 324, 3.4 L 300 324, CX CECR 15 339, CX CECR 1 340.

### 3.3 Use Case: Predict CVD in Diabetes Mellitus patients using Patient Trajectories

#### 3.3.1. Chosen parameters

To know which clinical variables are of interest when it comes to relating CVD with diabetes, an extensive search on risk prediction models was made. Table 2 shows the variables that appeared somehow in the risk prediction models proposed in the reviewed studies. The most used parameters in Table 2 would have been the parameters to ideally consider but not all of them were available in the EHR. Some of them, such as height, weight or blood pressure, are usually annotated in free text during anamnesis. Sex is a relevant factor for CVD since its incidence rate is 4 times higher in diabetic versus non-diabetic women, whereas this ratio is 2.5 in men [12]. This difference is due to the different HDL levels in both sexes, having women usually higher, and so more protective, levels. Diabetes usually decreases HDL levels, causing to lose this advantage.

**Table 2:** Variables included in each of the cited studies. Total column shows how many times each variable has been used in risk prediction models.

| Variable | [19] | [11] | [12] | [20] | [21] | [22] | [23] | [24] | [25] | [13] | Total |
| --- | --- | --- | --- | --- | --- | --- | --- | --- | --- | --- | --- |
| HDL Cholesterol | ☒ | ☒ | ☒ | ☒ | ☒ |  | ☒ | ☒ | ☒ | ☒ | 9 |
| Systolic, diastolic pressure or hypertension | ☒ | ☒ | ☒ | ☒ | ☒ |  |  | ☒ | ☒ | ☒ | 8 |
| Total Cholesterol (TC) | ☒ |  | ☒ | ☒ | ☒ |  | ☒ | ☒ | ☒ | ☒ | 8 |
| Sex |  | ☒ | ☒ |  | ☒ | ☒ | ☒ |  |  | ☒ | 6 |
| Smoking | ☒ |  | ☒ | ☒ | ☒ |  |  | ☒ |  | ☒ | 6 |
| Glycosylate haemoglobin (HbA1c) | ☒ |  | ☒ |  | ☒ | ☒ | ☒ | ☒ |  |  | 6 |
| Age |  | ☒ |  | ☒ | ☒ |  |  | ☒ |  | ☒ | 5 |
| BMI | ☒ | ☒ |  | ☒ |  | ☒ |  |  |  | ☒ | 5 |
| Diabetes time length | ☒ |  | ☒ |  | ☒ |  |  |  |  | ☒ | 4 |
| LDL Cholesterol | ☒ |  | ☒ |  |  |  |  |  | ☒ | ☒ | 4 |
| Creatinine |  |  |  | ☒ | ☒ | ☒ | ☒ |  |  |  | 4 |
| Age at diagnosis | ☒ |  | ☒ |  |  |  | ☒ |  |  |  | 3 |
| Tryglyceride | ☒ | ☒ |  |  |  |  |  |  |  | ☒ | 3 |
| Ethnic |  |  | ☒ | ☒ |  |  |  |  |  |  | 2 |
| Familiar history of diabetes |  | ☒ |  |  |  |  | ☒ |  |  |  | 2 |
| Height | ☒ |  |  |  |  |  |  |  |  |  | 1 |
| Haemoglobin (Hb) |  |  |  |  |  | ☒ |  |  |  |  | 1 |
| Hips-Waist ratio |  |  |  | ☒ |  |  |  |  |  |  | 1 |
| Physical activity |  |  |  | ☒ |  |  |  |  |  |  | 1 |
| Coagulation factor 8 |  |  |  | ☒ |  |  |  |  |  |  | 1 |
| Previous CVD |  |  |  |  |  | ☒ |  |  |  |  | 1 |
| Retinopathies |  |  |  |  |  |  | ☒ |  |  |  | 1 |

Although diagnostics and consultations are not directly used by the prediction models reported in the literature, we included them as observations of the patient trajectories. Moreover, we have access to the information about the place where the diagnosis was made (hospitalization, DH, or emergency room, DE). This was also included in the patient trajectories following the work of Jensen et al. [1].

Finally, the selection of clinical variables to be considered is (1) sex, (2) diagnostics (ICD-9-CM), (3) outpatient consultations, (4) total cholesterol, (5) HDL, (6) creatinine and (7) glycated haemoglobin. In addition, some nephrological diseases can increase the chances of having CVD in patients with diabetes [12], so ICD- 9 codes from chapter 10 will be specifically considered for the delta function. We specified the similarity of these parameters in different delta matrices that will be used by the delta function. We defined a total of 12 different scoring matrices, one for each type of observation, that can be seen already optimized in Figure 3. There is an explained example of how these scoring matrices are used together with the modified Smith-Waterman algorithm in Figure A.2.

#### 3.3.2. Experiments

The main experiment we performed to optimize the LPTA for the use case aimed to find the best weight for each one of the defined parameters, so its output is the scoring matrices in Figure 3. As the number of parameters is large, our strategy was the following: (1) fix a negative value both for those parameters not directly related to a CVD development (e.g protective levels of HDL) and for cases where different parameters are being compared. (e.g one diagnosis event and one laboratory test), (2) set the rest of parameters to 0, (3) evaluate the performance of the algorithm when varying each parameter when they take different values 1, 3, 5, 7, 9, (4) for each parameter, the lowest value with the highest performance was preferred. After fixing these values, we run a final experiment in order to determine which number of patients (N) for the classification method gives the best results: 1, 2, 5, 10, 15, 25, 40, 60, 80, or 100.

#### 3.3.3. Evaluation

The PTs of the CVD validation patients were cut before one of the CVD diagnostics appeared (i.e. ICD-9- CM codes 410,411,412,413,414, 427.1,427.3,427.4, 427.5, 428, 429.2, 440.xx, 440.23, 440.24, and 441), therefore some of the PTs had to be removed as the CVD diagnosis was the first event recorded in their EHR and there were not more events in the PT to make the alignment. For evaluating the generalizability of the results, a cross-validation with 10 folds was made. Due to the high computational cost of the experiments, a training set of 800 patients and a validation set of 200 patients were randomly selected for each experiment from the corresponding cross-validation partition, as shown in Figure 2.

**Figure 2:**
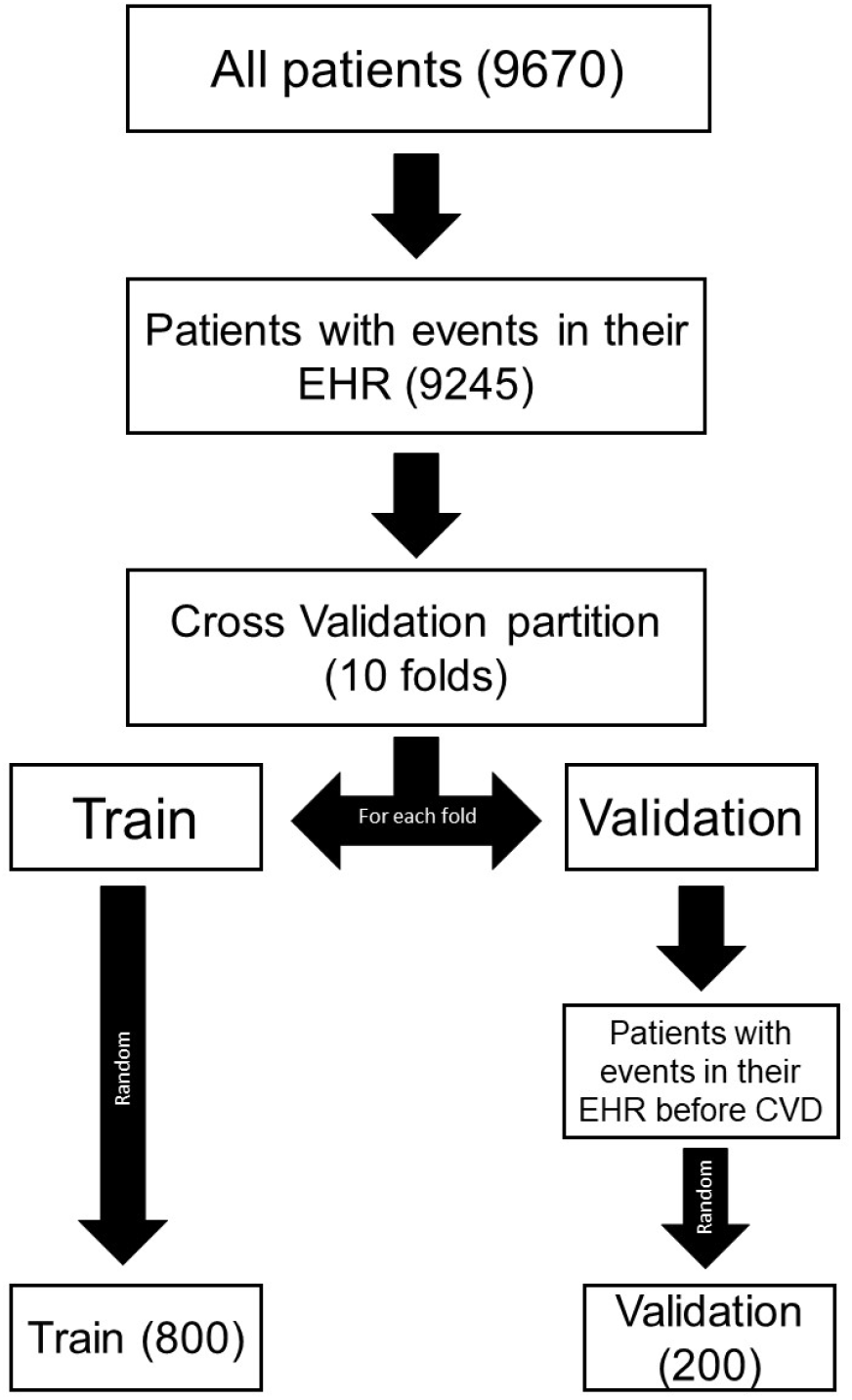
Obtainment process of the train and validation sets for the experiments. PTs of the test set patients are cut before the CVD appears.

Precision, Recall and Specificity of the results were measured in each experiment. Precision, also called positive predictive value, indicates how many of those selected as CVD patients by the algorithm are really CVD patients. Recall or Sensitivity indicates how many of those who are CVD patients are selected by the algorithm. Specificity indicates how many of those who are not CVD patients are correctly identified as non-CVD patients by the algorithm. Generally, there is a compromise between specificity and recall so the greater the specificity, the lower the recall and vice versa. Since the algorithm is to be applied in a clinical setting as secondary screening, it is advisable to have a conservative perspective, which is why a high recall is preferred over high specificity.

## 4. Results

After iterating with several values, the best results of the matrices are those shown in Figure 3. The parameters of the delta matrices with the highest weight for predicting CVD-development in diabetes mellitus were (1) the exact match of the ICD-9 code, (2) diagnostics of the cardiology chapter, (3) cardiology consultations, (4) very high total cholesterol, (5) high HbA1c, (6) high HDL in case of women and (7) coincidence in the time parameters, therefore they are the most related to the development of a CVD in patients with diabetes.

**Figure 3:**
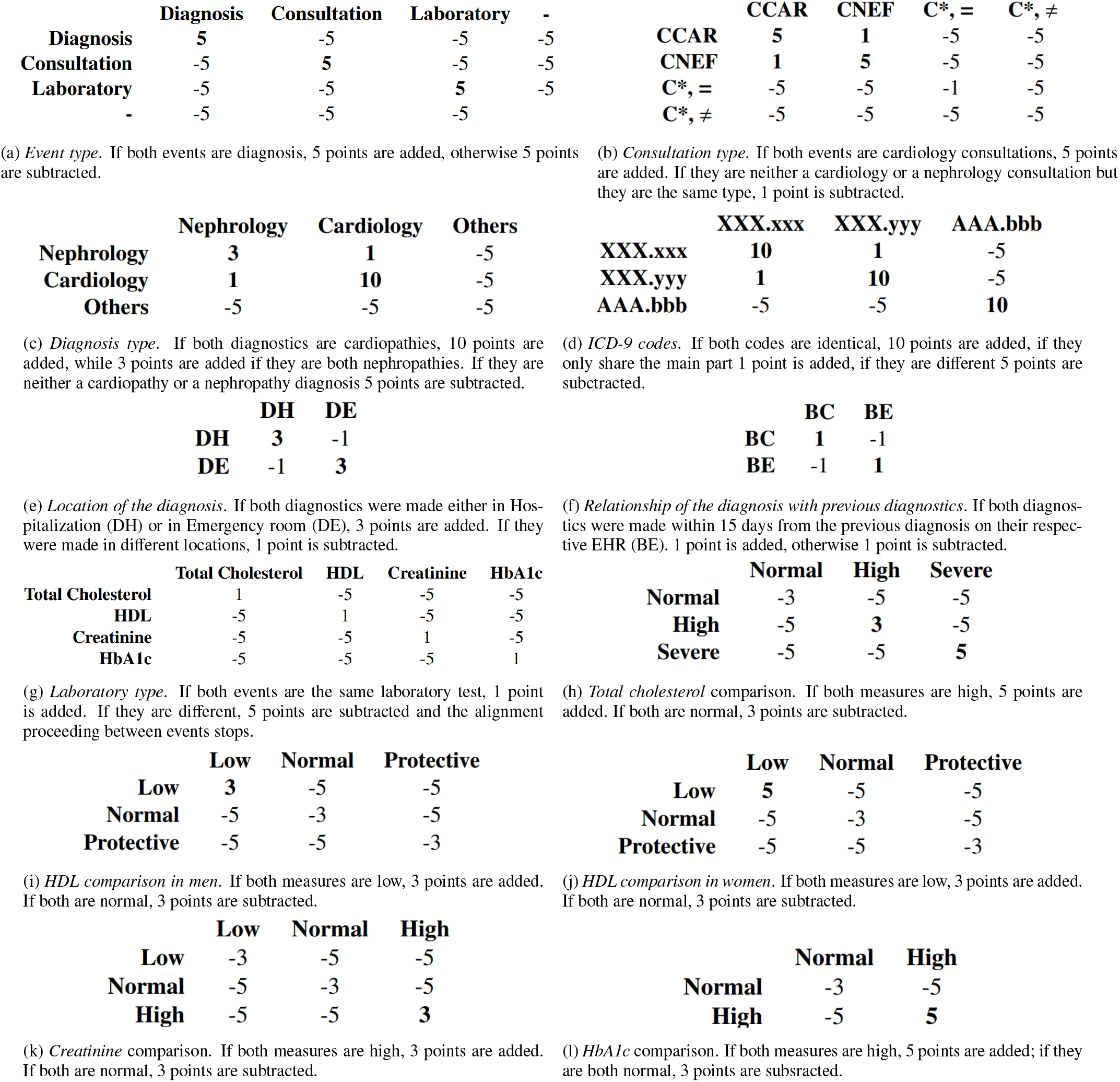
Alignment scoring matrices optimized to our diabetes use case. (3a) is the main matrix, followed by (3b), (3c) and (3g) depending on the event type. Matrices (3d), (3e) and (3f) will be used if both events are diagnsoses, while (3h), (3i), (3j), (3k) amd (3l) will be the ones used if both events are laboratory tests. When evaluating the similarity of time parameters, five points would be added if they are similar while a point would be subtracted if they are not similar, considered as similar time frames time differences of less than 15 days, as explained in section 3.2.

Once the scoring matrices were fixed, an extra experiment was performed to choose the best number of patients which condition is consulted for the classification method and its results can be seen in Figure 4. When N was set to 5, which represents imputing the CVD condition if at least 1 out of the 5 most similar patients has developed a CVD, LPTA-based classification method obtained its best results (positive predictive value of 0.33, recall of 0.72 and specificity of 0.38).

**Figure 4:**
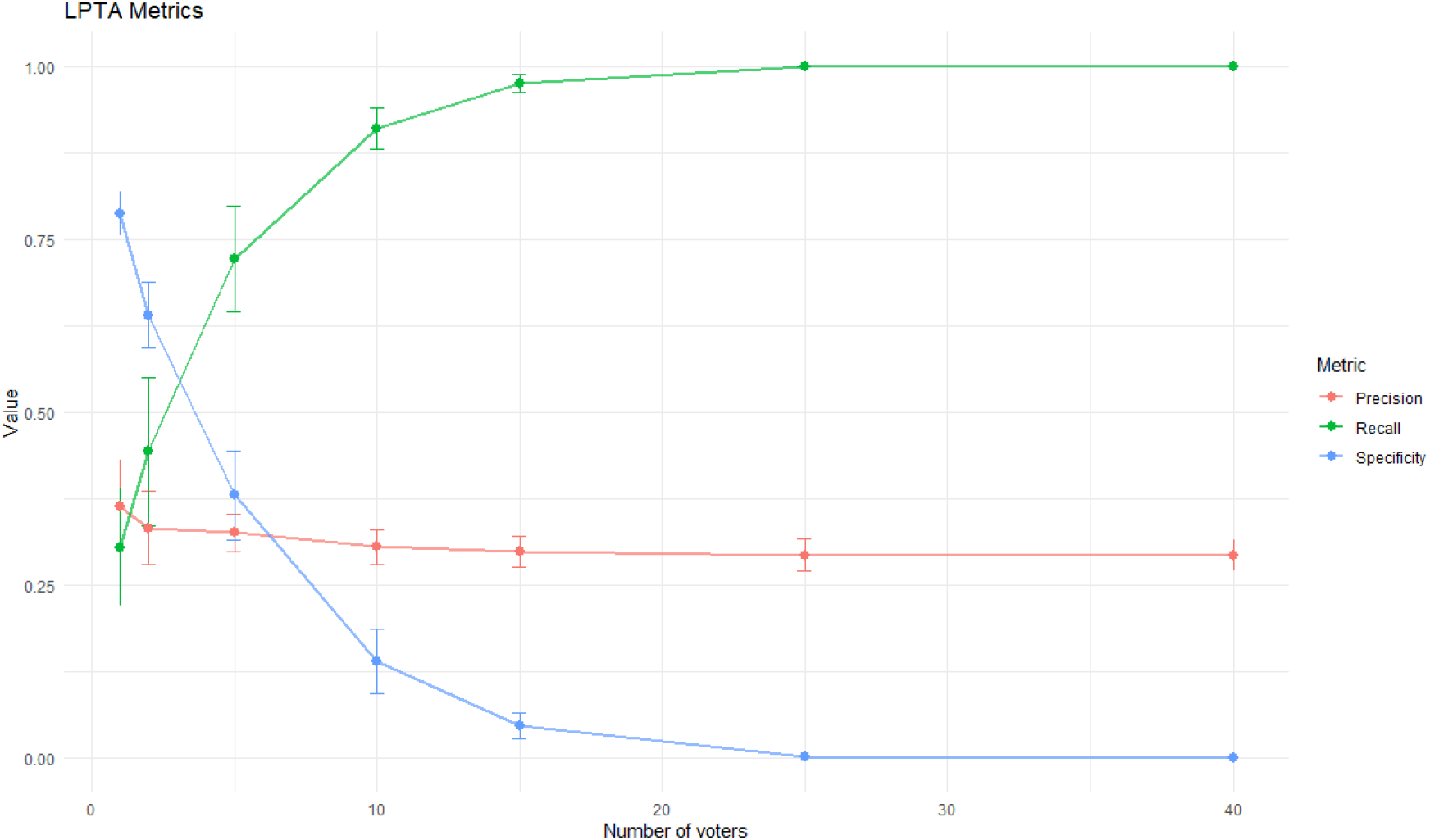
LPTA results according to the number (N) of most similar patients which condition is consulted to assign the development of the condition to the query patient. This figure shows the compromise between sensitivity and specificity mentioned in Section 3.3.3, as one converges to 1 while the other converges to 0.

## 5. Discussion

Although some studies about patient trajectories analytics have focused their attention on the sequential representation of patients’ health records, to the best of our knowledge this is the first study to predict potential morbidities in patients based on local similarities of PTs. This simple but powerful operation has proved to be useful as a secondary screening method of patients with diabetes mellitus based on patient trajectories. Solving this task using patient trajectories instead of the classic multiparametric representations may draw on the temporal relationships of the observations. The other great contribution of this work is that it is not necessary to generate aggregate PT from the reference dataset, as is done in the works reviewed in Section 1.1. In this work, the similarity measure is calculated for each of the available PTs, so that the comparisons made are more accurate and there is no loss of information.

A formal definition for patient trajectories has been proposed. PTs can be used not only for local alignment but also for dealing with different issues, such as EHR-data visualization or detecting patterns in data, as we have seen in Section 1.1. It would not be difficult to add new information as convenient, such as Patient-Reported Outcomes (PROs) or Quality-adjusted life year (QALY), in order to evaluate different therapies or disease trajectories. It could also be added any other clinical information such as secondary diagnostics or DRG codes to have more relevant information included in the PTs.

The LPTA algorithm has proved to be useful when finding similar regions in PTs. If these common regions are sufficiently similar, the condition of one of the patients can be imputed to the other one, as it has been done in our use case. Generally speaking, although the amount of data available for each patient may be different, as there are persons that visit the hospital more frequently than others, significant local similarities can be detected by the LPTA algorithm. Moreover, normalizing the similarity score by the number of observations in the trajectory of the patient reduces the influence of the PT length.

We were concerned that the length of the PTs was a determining factor in the performance of the algorithm, thinking that the shorter the PTs, the less information the algorithm would have to evaluate. Previous experiments were carried out and it was finally determined that, although the length of the PT slightly affects the algorithm, it is not enough to justify the elimination of the study of patients who do not have enough information in their EHR. The main use we see for LPTA is screening, so it should be able to be applied to as many patients as possible.

Several applications of the proposed algorithm arise. While the LPTA has proved useful for screening in our case study, for other problems it could also be useful for diagnosis or prognosis. It could be also used for detecting similarities of PTs for further understanding of rare diseases, detecting similarities in different population groups or predicting whether a patient could benefit from a particular treatment. The algorithm can be easily adapted to different datasets since the variables available can change from one use case to another.

### 5.1. Limitations

One of the main limitations of this algorithm is its temporal cost, similar to the Smith-Waterman’s computational cost, (*O*(*n*^2^)), with n the mean number of events in both sequences. This large temporal cost is also reported in Sha et al. work [9], being up to six times higher than other similarity measures such as the Jac- card similarity coefficient. A Big Data technology to speed up the computation of LPTA is already being developed [18]. Although this problem is easily adaptable to other diseases, dealing with high-dimensional data can be complex. The more variables are included, the larger the scoring matrices will be. However, as stated, the matrices are divided into sub-matrices according to sub-domains, allowing the reuse of some of them in different problems (e.g the score associated with a visit to a traumatology consultation may be the same whether the development of heart disease or nephropathy is being predicted).

In addition, although we had more than 20 parameters to evaluate the similarity, some parameters considered as important in risk prediction models such as BMI or blood pressure were not included in the algorithm as they were not available in our dataset. The inclusion of these parameters, in addition to others such as medication and race, may improve the results of the algorithm. Finally, there is an implicit limitation regarding the temporal development of the disease. Some of the patients that were labelled as non-CVD-developers when the dataset was extracted may have developed a CVD afterwards, so they should not be considered as errors from the classifier if classified as CVD-developers.

The search for values for the matrices performed in the optimization experiment was not continuous, so the resulting values may not be optimal. In addition, as some values were pre-set and not optimized, it may also have led to sub-optimal results for the other parameters.

There is an implicit problem with the number of false positives, whose probability of occurrence increases as the number of cases analyzed increases. Final specificity and positive predictive value may not be the desired, but recall is high (0.72). The proposed algorithm is presented as a secondary screening method, so a high recall and an acceptable specificity is wanted, which have been achieved in the experiments. Another work that was based on the alignment of history and used a Smith-Waterman based similarity measure [9] also achieved similar results, with a specificity around 0.7 and a recall around 0.6. Although these results seem limited compared to those obtainable by other methods of the state-of-the-art like Machine Learning (ML), the LPTA offers the advantage of being able to recover which part of the trajectory caused the classification, so it is not a black box like what ML can be. By showing the physician the part of maximum similarity with the most similar reference patient’s PT, he or she can easily understand which parts of the patient’s clinical history most determine his or her condition.

## 6. Conclusions

This work has led to the following contributions: (1) a formal definition of patient trajectory based on heterogeneous sequences of multi-scale data over time, (2) a dynamic programming methodology to identify local alignments in patient trajectories with customized matrices, and (3) a specific LPTA-based classification method to predict the development of CVD in patients with diabetes mellitus that achieved a precision of 0.33, a recall of 0.72 and a specificity of 0.38. The most prevalent conditions in the local chunks of PTs predicting cardiovascular diseases in diabetes patients included cardiology diagnosis and consultations, serious levels of total cholesterol, and high HbA1c. The proposed PT definition has been tested in a specific CVD use case, but it could be generalized to further domains, adapting it to include additional variables and cost matrices without changing the algorithm. To our knowledge this is the first methodology where patient trajectories have been modelled as a sequence of multi-scale data aiming to their local alignment through a dynamic programming algorithm to identify future morbidities. This approach is able to evaluate the similarity in local chunks of trajectories being robust to heterogeneous global trajectories in terms of length and disease temporal patterns spread along the patient life.

## 7. Ethics approval and consent to participate

Approved by the Ethical Committee of Hospital Universitario y Politecnico La Fe under the Project ’’Modelos y tecnicas de simulación para identificar factores asociados a la diabetes” presented by Dr. Bernardo Valdivieso with code: 2015/0458.

## Data Availability

The data is not available to the public.

## Appendix A. Supplementary material

**Figure A.1:**
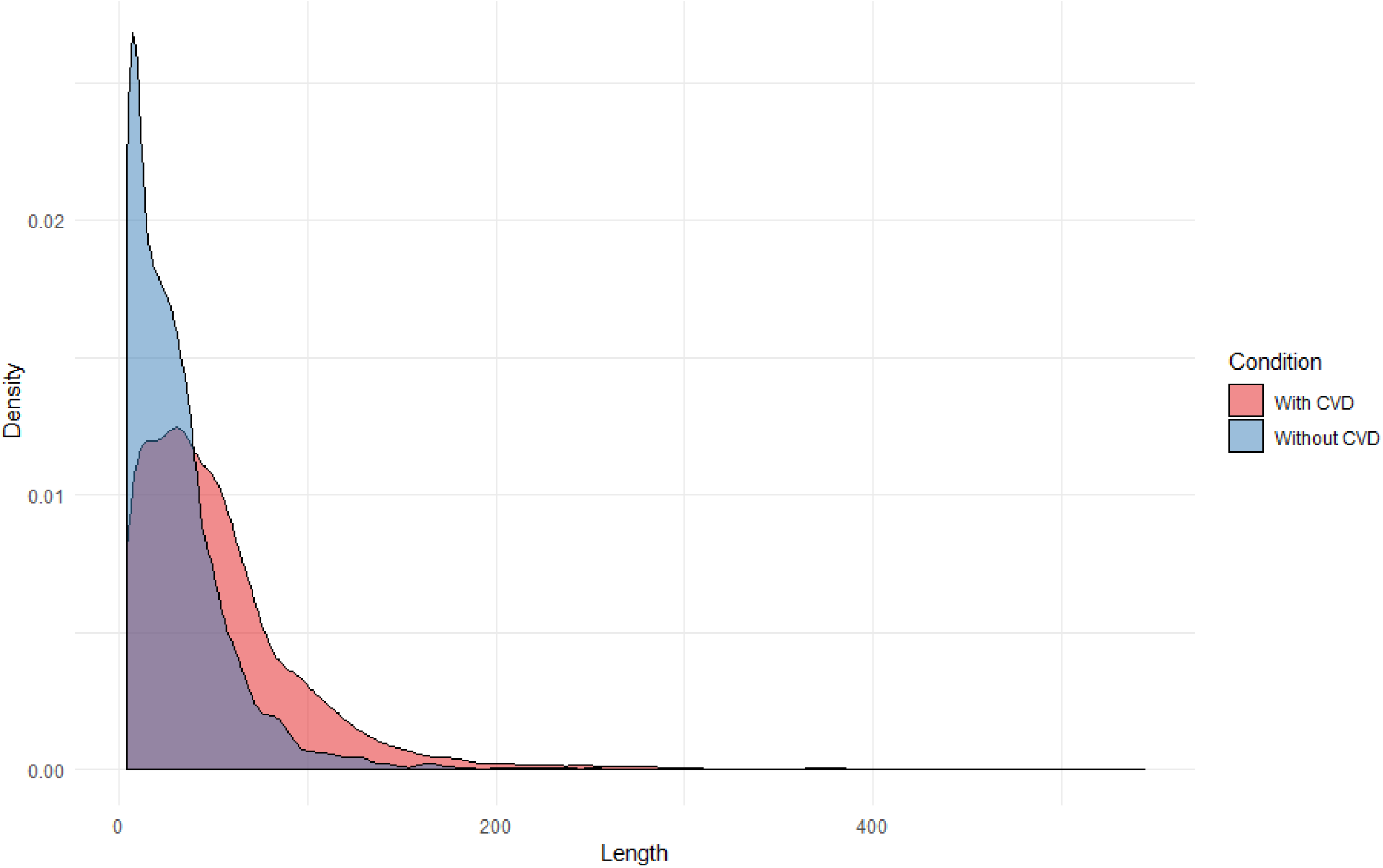
Distribution of the number of events per patient in their EHR. CVD patients have longer trajectories, while most of the non-CVD patients have less than 10 observations.

**Function Appendix A.1:**
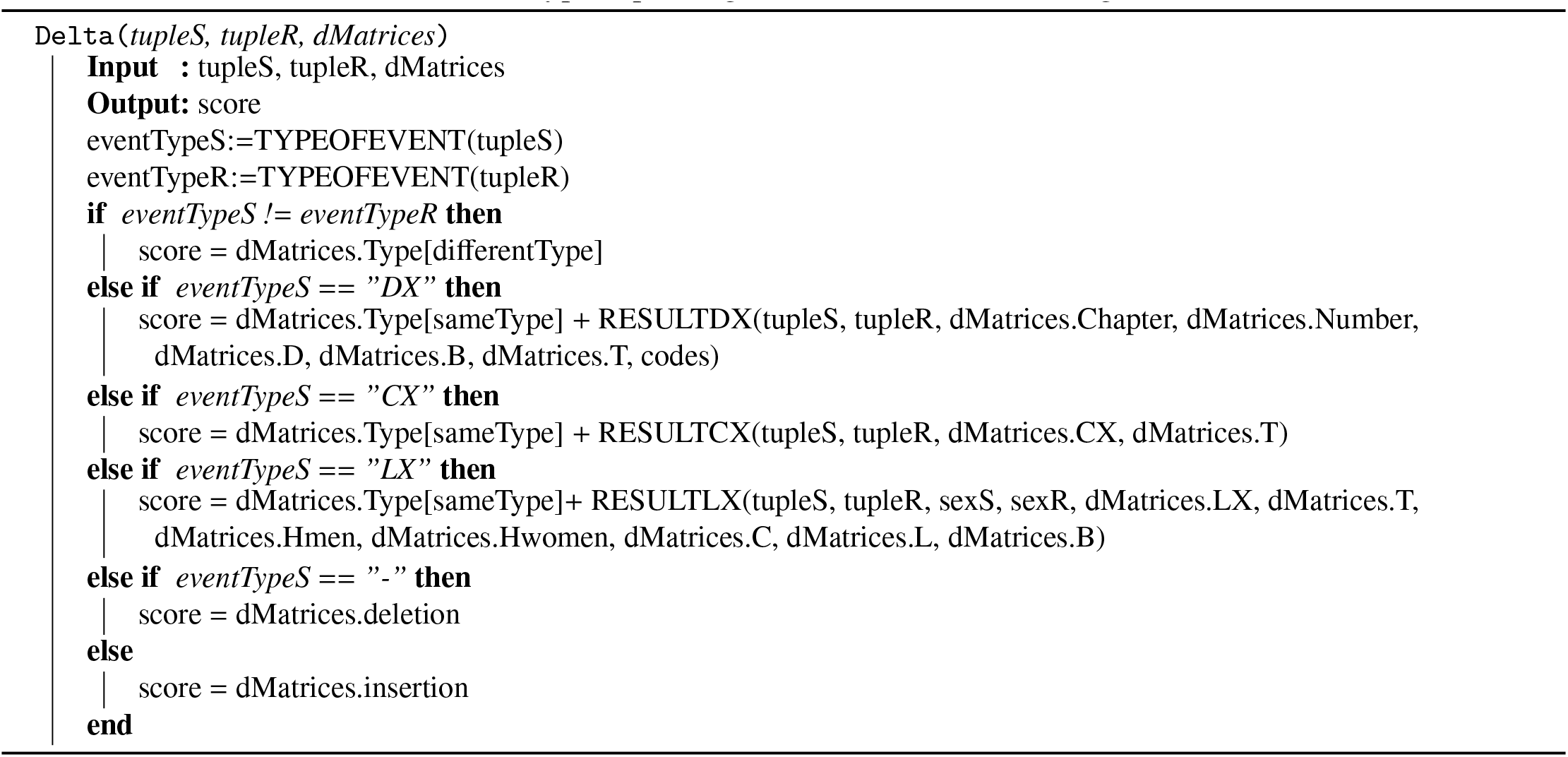
Delta scoring function. tupleS is an observation in a query patient trajectory and tupleR is an observation in a reference patient trajectory. TYPEOFEVENT is a function which output is the type of event that the tuple is: CX for consultations, DX for diagnosis and LX for laboratory tests. RESULTDX, RESULTCX (Function Appendix A.2) and RESULTLX are functions which output is the similarity score between two observations of the same type depending on the values of the scoring matrices.

**Function Appendix A.2:**
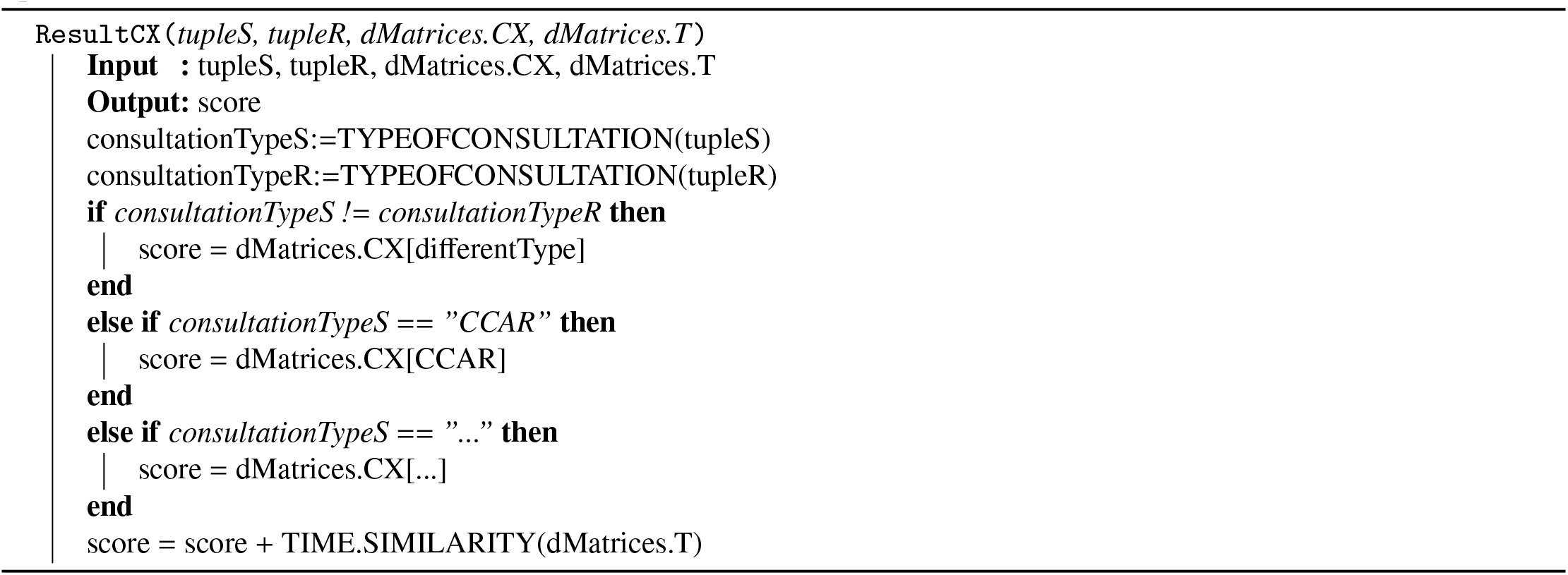
ResultCX. For a further understanding of how the scoring functions work, RESULTCX is shown. In dMatrices.CX we have different scores depending on the consultation type. TIME.SIMILARITY will evaluate the similarity of available time parameters and will result in a score depending on it.

**Figure A.2:**
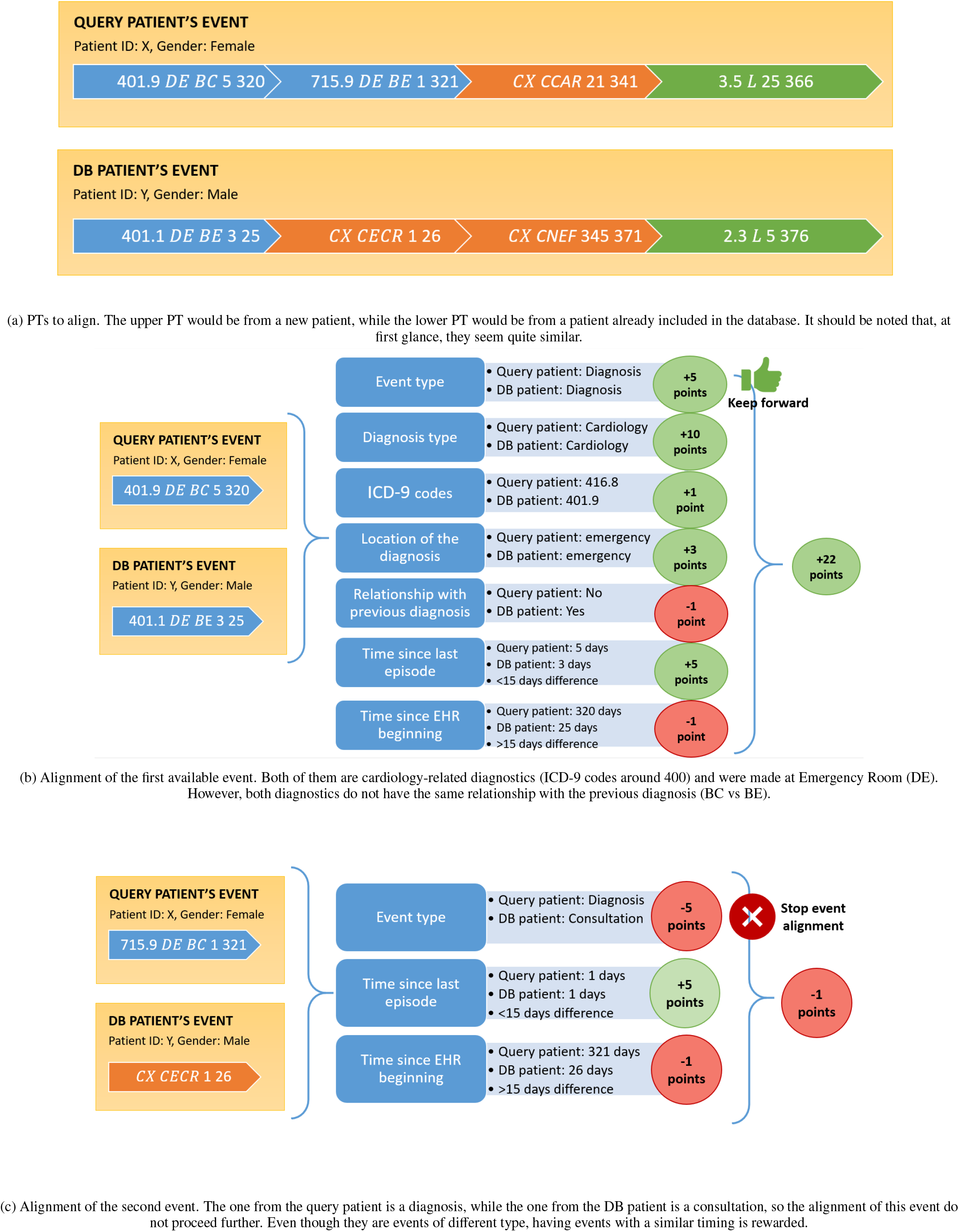
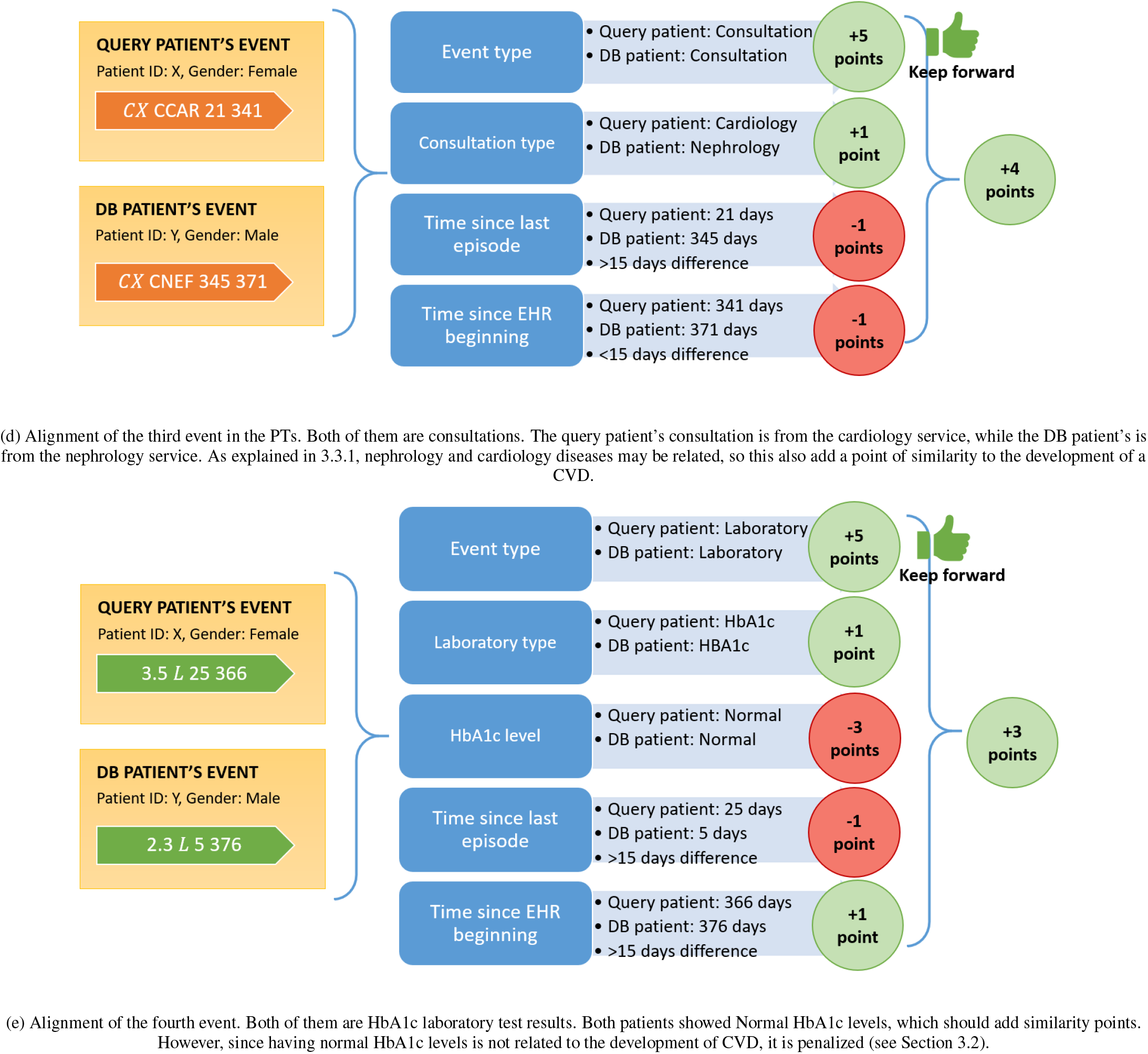
Example of an alignment between a new query patient’s PT and a PT from a patient in the database. This alignment is done by substitution or match, not by insertion or deletion (see Section 1.2), so it might not be the optimum. The final similarity score between the PTs in Figure A.2a would be of 27 points (22 − 1 + 4 + 3 = 27). The normalized score (see Section 3.1) would be of 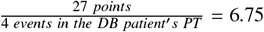

